# Heart Rate Variability as a Prospective Predictor of Early COVID-19 Symptoms

**DOI:** 10.1101/2021.07.02.21259891

**Authors:** Alexey Ponomarev, Konstantin Tyapochkin, Ekaterina Surkova, Evgeniya Smorodnikova, Pavel Pravdin

## Abstract

Heart rate variability (HRV) is the fluctuation in the time interval between consecutive heartbeats, the measurement of which is a non-invasive method for assessing the autonomic status. The autonomic nervous system plays an important role in physiological situations, and in various pathological processes such as in cardiovascular diseases and viral infections. This study examined the cardiac autonomic responses, as measured by HRV before, after, and during coronavirus disease. In this study, we used beat interval data extracted from the Welltory app from 14 eligible subjects (9 men and 5 women) with a mean age (SD) of 44 (8.7) years. HRV analysis was performed through an assessment of time-domain indices (SDNN and RMSSD). Group analysis did not reveal any statistical difference between HRV metrics before, during, and after COVID-19. However, HRV at the individual level showed a statistically significant individual change during COVID-19 in some users. These data further support the usefulness of using individual-level HRV tracking for the detection of early diseases inclusive of COVID-19.

## Introduction

The coronavirus disease 2019 (COVID-19) is a highly contagious and pathogenic viral infection caused by the novel coronavirus, severe acute respiratory syndrome coronavirus 2 (SARS-CoV-2). COVID-19 triggered a global pandemic that has created serious challenges for medical systems and civilian communities, resulting in enormous human loss worldwide throughout 2020 and 2021. (Hu et al., 2021; Taleghani & Taghipour, 2021). To date, the World Health Organizations (WHO) and health care systems in different countries around the world are struggling to control the spread of COVID-19. A fast and accurate self-test tool for early detection of COVID-19 can be helpful in improving responses to this disease.

Heart rate variability (HRV) is the fluctuation in the time interval between consecutive heartbeats called interbeat interval (R-R interval; Montano et al., 1994). This indicator is associated with the regulatory and homeostatic functions of the autonomic nervous system (ANS), including cardiac parasympathetic nervous activity (cPNA) and cardiac sympathetic nervous activity (cSNA). An optimal level of HRV is associated with health and self-regulatory capacity, adaptability, and resilience. HRV is of interest as a wide range of diseases are associated with decreased variability, including diabetes, cardiovascular disease and psychiatric disorders (An et al., 2014; Benichou et al., 2018; The et al., 2020). HRV indicators are often used to determine the risk of developing certain pathological conditions. For example, studies have shown an association between low HRV and mortality in patients with coronary artery disease and chronic heart failure (Huikuri & Stein, 2013). Low HRV also predicts the risk of heart morbidity and mortality in seemingly healthy people (Hillebrand et al., 2013). Interestingly, higher HRV does not always indicate obvious protection, as high HRV poses the risk of atrioventricular block, sick sinus syndrome, and atrial fibrillation (Fu et al., 2007).

HRV analysis can be conducted in the time domain (the standard deviation of all R–R intervals and the root mean square of successive standard deviation), frequency domain (different spectral profiles of the oscillatory components), and by using non-linear analyses (Shaffer & Ginsberg, 2017). Use of HRV has substantially increased in recent decades in research and clinical treatment applications (Kim et al., 2018).

COVID-19, along with other viral infections, is associated with several physiological changes that can be monitored using wearable sensors (Quer et al., 2021). For example, it became known that hypertension or coronary heart disease is present in 38% of COVID-19 patients (Zhou et al., 2020). Since a significant correlation of cardiovascular diseases with changes in the HRV indicators was previously shown, the measurement of these indicators can be used for early detection of COVID-19. Several studies have already shown the usefulness of using indicators derived from heart rhythm such as heart rate (HR), heart rate variability (HRV), resting heart rate (RHR), and respiration rate (RR) for COVID-19 early detection (Natarajan et al., 2020; Tanwar et al., 2020). Users of wearable devices could be notified when changes in their HRV metrics coincide with those associated with COVID-19 and then alerted of a potential SARS-CoV-2 infection before symptoms become severe. This alert can facilitate early initiation of appropriate therapy and reduce the development of respiratory and cardiovascular complications.

In this article we tested the hypothesis that HRV could serve as an early potential marker of COVID-19 infection.

## Materials and methods

### Study design

This study was conducted without the active participation of the research participants. Upon downloading the Welltory app, users provide informed consent for their anonymized data to be used by the company for internal research purposes if such research can help provide users with better services or improve the app’s functionality. This policy is described in the company’s Terms of Use, which the app’s users actively consent to.

The data was collected between March 2020 and March 2021, during the period marked by the greatest spread of the SARS-Cov-2 pandemic around the world.

In this study, we used interbeat interval data extracted from the Welltory app. These data were collected through photoplethysmography (PPG) technology with smartphone cameras (Tyapochkin et al., 2019), wrist-worn smartwatches, and wrist-worn bands synchronized with the Welltory app. Age, sex, weight, and height were also extracted from the Welltory app.

For each user, a set of the following parameters was obtained: SDNN, MEDSD, and RMSSD. SDNN is the standard deviation of NN intervals. To exclude the influence of cardiac automatism, we used a corrected SDNN based on the human heart rate (Natarajan et al., 2020). MEDSD is the median of the difference of consecutive intervals. RMSSD (“root mean square of successive differences”) is the square root of the squares of the successive differences between NN intervals, essentially the average change in interval between beats.

To exclude the influence of cardiac automatism, we used a corrected RMSSD based on the human heart rate (de Geus et al., 2019). We used the exponential model for a correction of the HRV markers (de Geus et al., 2019).

People with the COVID-19 were invited to participate in this study by tracking their symptoms, heart rate variability, and other data from wearables such as Apple Watch, Garmin, or Fitbit with the Welltory app. Participants added a COVID-19 tag to their measurements. The following tags were found: “covid”, “coron”, “virus”.

We announced the launch of our data collection and research project on the Welltory platform in April 2020 (https://github.com/Welltory/hrv-covid19). Unfortunately, the App Store policy blocked our app due to the mention of COVID-19. Thus, we were forced to remove any mention of COVID-19 from the Welltory app. Moreover, users could only create tags related to COVID-19 by themselves. This is clearly shown in Figure 1.

**Figure 1.**
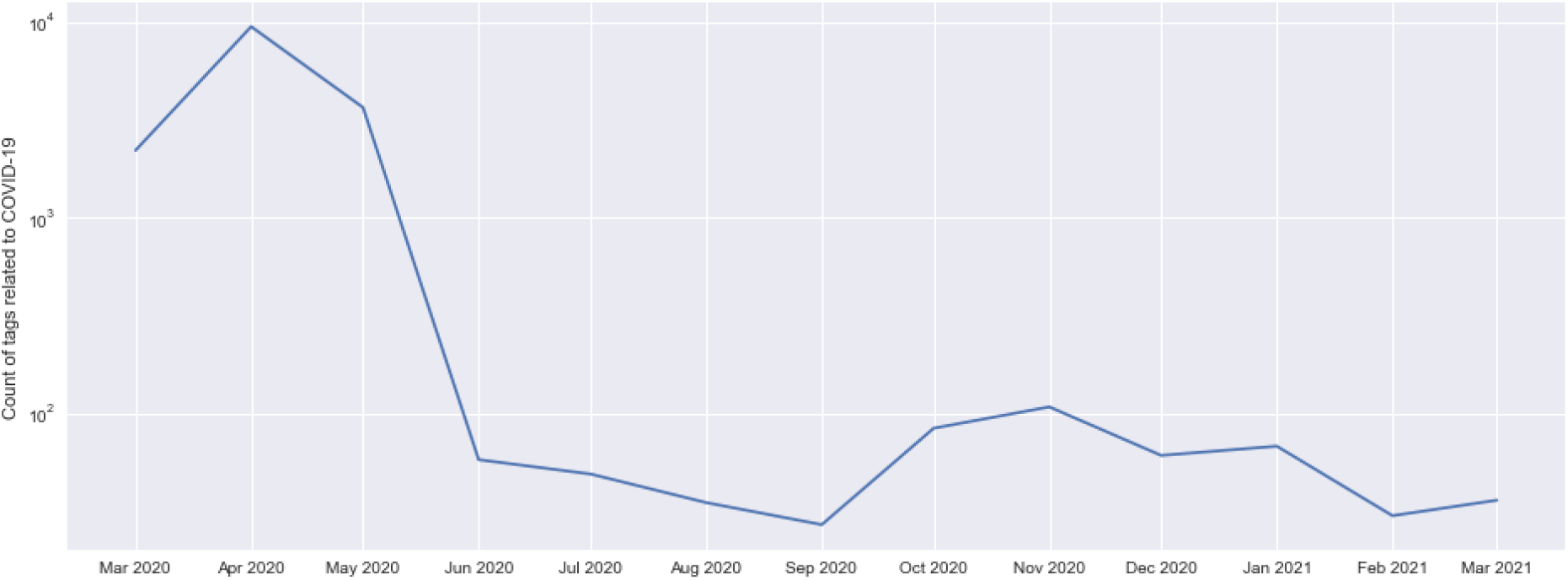
User activity in tagging related to COVID-19 in the Welltory App from March 2020 to March 2021.

We used data from people who used many COVID-related tags for the analysis. The measurement interval between adjacent tags was to be no more than three days. All measurements were divided into three groups: before vs. COVID-19; COVID-19 vs. after; and (before + after) vs. COVID-19.

### Statistical analysis

Paired t-tests were used to compare data before and during the experience of COVID-19 symptoms for all users. The Mann-Whitney test with two one-sided alternatives was used to compare data before and during the experience of COVID-19 symptoms in individual users. The significance level for all tests was set to 5% (α = 0.05).

## Results

We found 15880 measurements with tags related to COVID-19. Users were most active in tagging the first wave of the epidemic (from March to May of 2020; Figure 1).

We then selected users who had tagged the disease multiple times over an extended period (Table 1). 495 users who had three or more days of COVID-associated tags were found.

**Table 1.** Number of users with different periods associated with COVID-19 tags.

| Days with COVID-19 tags | Number of users |
| --- | --- |
| 1 | 9107 |
| 2 | 648 |
| 3 | 199 |
| 4 | 77 |
| 5 | 63 |
| 6-10 | 86 |
| 11-20 | 56 |
| 21-50 | 14 |
| 51-100 | 0 |

Next, we analyzed the duration of the tagged continuous episodes (Table 2). 224 users who had 5-15 days of continuous COVID-associated tags were found. Of these, 148 users had one series of COVID-associated tags without tags before and after the series.

**Table 2.** Number of users with different durations of the COVID-associated tag period.

| Duration of tagged episodes | Number of users |
| --- | --- |
| 1 | 27 |
| 2 | 45 |
| 3 | 79 |
| 4 | 83 |
| 5-15 | 224 |
| 16-20 | 19 |
| 21-50 | 16 |
| 51-100 | 2 |
| 101-1000 | 0 |

We then analyzed all measurements for the selected users, starting one month before the first COVID tag in the series. For each day and each user, we used only one measurement for analysis, which was at the time of day when this user most often took measurements. We formed the cohort by including only those users who had more than 5 high quality measurements in the period before and during the COVID-19 tag in the analysis. The final cohort included 9 men and 5 women, mean age 44 ± 8.7 years, height 174 ± 12.9 cm, weight 80 ± 18.0 kg.

We compared corrected HRV indicators (SDNN and RMSSD) in three groups of measurements: before vs. COVID-19, COVID-19 vs. after, and before + after vs. COVID-19 (Fig. 2)

**Figure 2.**
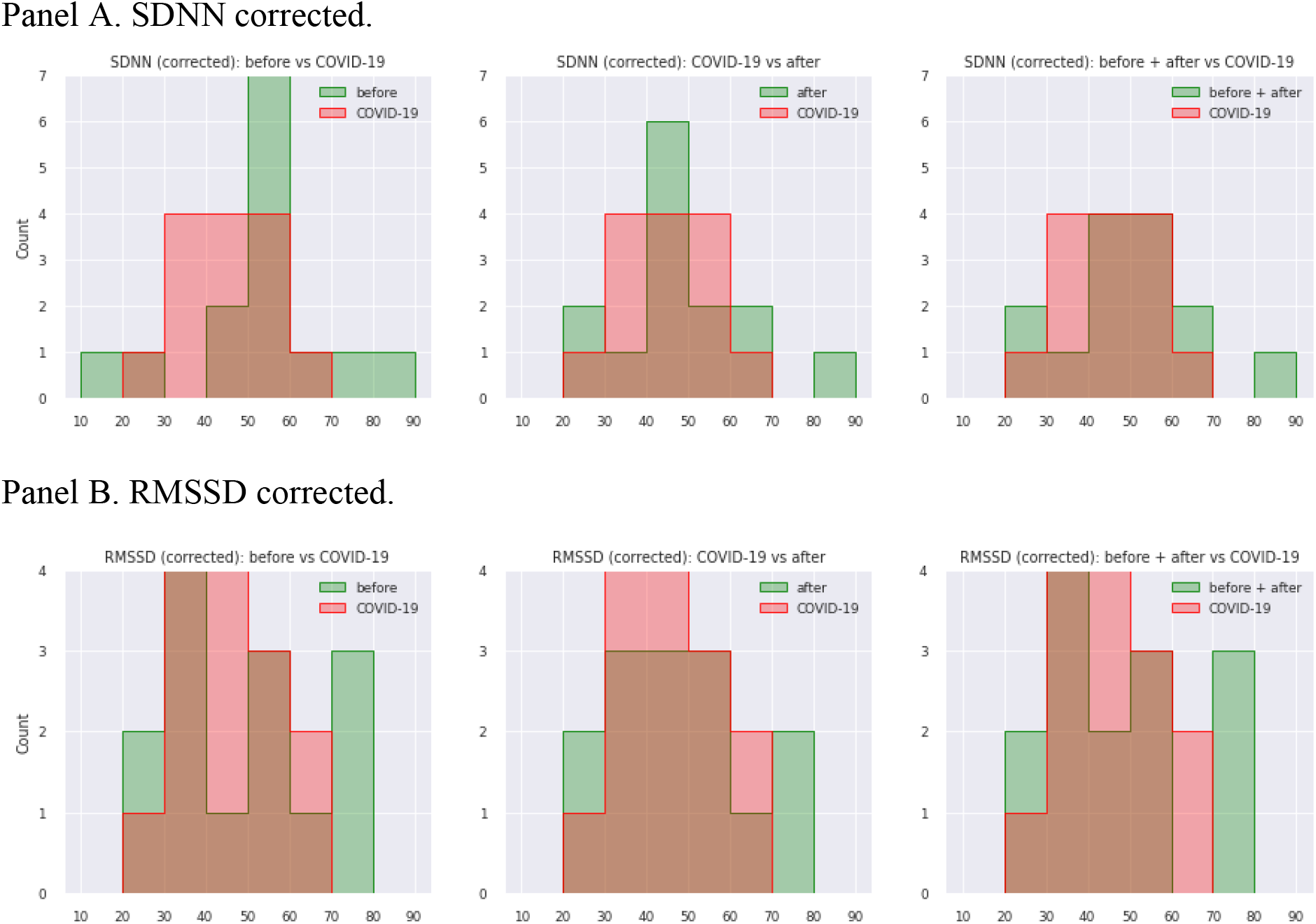
Histograms of HRV markers for three groups of measurements: before vs COVID-19, COVID-19 vs after, and before + after vs COVID-19. Panel A - SDNN corrected, Panel B - RMSSD corrected.

In our study, no statistically significant interaction was found between the HRV indicators before, during, and after COVID-19 illness in our cohort. When comparing periods before, during, and after COVID-19 illness in individual users, we found statistically significant differences in SDDN and RMSSD for some users (Table 3). The SDNN increased during disease for three users, while for one user it decreased (Table 3). The RMSSD was statistically significantly reduced for one user during COVID-19 illness (Table 3).

**Table 3.** SDNN and RMSSD corrected comparisons for individual users between two groups of measurements: before COVID-tag and during COVID-tag.

| User | SDNN p-value |  | RMSSD p-value |  |
| --- | --- | --- | --- | --- |
|  | SDNN before COVID-tag is **greater** than SDNN during COVID-tag | SDNN before COVID-tag is **less** than SDNN during COVID-tag | RMSSD before COVID-tag is **greater** than RMSSD during COVID-tag | RMSSD before COVID-tag is **less** than RMSSD during COVID-tag |
| 1 | .054099 | .952475 | .318301 | .703831 |
| 2 | .056388 | .948937 | .304204 | .712650 |
| 3 | **.029832** | .974669 | .652053 | .374560 |
| 4 | .985581 | **.017724** | .946119 | .063523 |
| 5 | .536368 | .536368 | .536368 | .536368 |
| 6 | **.044487** | .962169 | .080986 | .929767 |
| 7 | .402662 | .624333 | .096333 | .915147 |
| 8 | **.000811** | .999354 | .196651 | .821033 |
| 9 | .607904 | .463632 | .463632 | .607904 |
| 10 | .564504 | .500000 | .096938 | .928074 |
| 11 | .695796 | .357197 | .863873 | .170675 |
| 12 | .758797 | .271335 | .663292 | .371530 |
| 13 | .545099 | .500000 | .285547 | .751662 |
| 14 | .079498 | .937107 | **.011732** | .991536 |

*Note:* *p*-values are given for one-sided hypotheses regarding the SDNN increases or decreases. Statistically significant differences are highlighted in bold.

## Discussion

A wide range of available ambulatory medical and fitness devices use sensors to detect heartbeats. Such sensors are used for auscultation of the heart (sound), blood pressure measurements (pressure or oscillographic), pulse oximetry (optical), photoplethysmography (optical), and electrocardiogram (ECG) measurements. Personal wearable sensors — including watches, rings, clothing, earphones, and more — that allow for a person’s unique changes in the HR and HRV to be continuously monitored and benchmarked to themselves — rather than a population norm — may have great value; such sensors may improve wellness and prevent both mental and cardiovascular diseases. In the case of infectious diseases, such sensors can be used to detect an early stage of the disease, which in turn will allow for early treatment and a reduction in the number of complications.

In this study, we analyzed the relationship between indicators and the development of disease. We have published the anonymized data for researchers as multiple comma-separated values (CSV) files in Github (https://github.com/Welltory/hrv-covid19). Using this data, Tanwar and colleagues showed that the onset or worsening of COVID-19 symptoms had a higher probability if the HRV components displayed a consistent decline state (Tanwar et al., 2020). Natarajan and colleagues also found a decrease in HRV during COVID-19 illness in 2,745 participants with PCR positive tests (Natarajan et al., 2020). Hasty and colleagues revealed that 72 hours before the increase of C-reactive protein levels, there was a decrease in HRV by more than 40% in patients with COVID-19. An increase in C-reactive protein levels is used to track the inflammatory process and is associated with the progression of this disease (Hasty et al., 2021).

However, we found no statistically significant differences in the HRV markers (SDNN and RMSSD) between periods before, during, and after COVID-19 illness for our cohort. Our sample size was modest, which limits our statistical power and, consequently, the strength of our conclusions. We were not able to gather enough data as a result of the App Store’s blocking of the Welltory app for its mention of COVID-19. In our study, too few users had a history of measurements suitable for qualitative research, which would combine good quality measurements, the presence of tags, and a long series of tags.

While we did not find a significant and persistent difference in HRV responses among periods before, after, and during COVID-19 illness, we found a trend towards such differences for individual users. A review of literature suggests that differences in contextual factors such as age, health, psychological condition, and medications can impact an individual’s HRV, rendering the tracking of individualized responses even more important (Garakani et al., 2009; Hughes & Stoney, 2000). Therefore, further studies regarding the association of HRV metrics with COVID-19 in large groups of users unified in terms of age, gender, and health are required. The findings support the value of HRV tracking in objectively measuring an individual’s response to potentially stressful stimuli.

Our study suggests a possible effect of COVID-19 on HRV metrics, so that it can be considered in designing future studies. The ability to objectively assess individual physiological responses to various diseases in real-life settings at scale and at a fraction of the cost can help support early disease detection, an assessment of potential disease threats, and an assessment of various treatment benefits.

## Conclusions

Simple wearable sensors allow for monitoring of HRV during daily activities. Personalized HRV analysis at the individual level showed a trend in HRV changes during COVID-19, which further confirms the usefulness of using HRV tracking at the individual level to objectively measure the response to various conditions. This is promising for early detection of various diseases including COVID-19. However, significant challenges remain for fully understanding how best to measure HRV changes in order to extract data that can be useful to humans.

## Data Availability

Due to the nature of this research, participants of this study did not agree for their data to be shared publicly, so supporting data is not available.

## Abbreviations

ANS: autonomic nervous system
COVID-19: coronavirus disease 2019
cPNA: cardiac parasympathetic nervous activity
cSNA: cardiac sympathetic nervous activity
ECG: electrocardiogram
HR: heart rate
HRV: heart rate variability
PPG: photoplethysmography
RHR: resting heart rate
RR: respiration rate
RMSSD: root mean square of successive normal-to-normal interval differences
SARS-CoV-2: severe acute respiratory syndrome coronavirus 2
SDNN: standard deviation of normal-to-normal intervals

## Acknowledgements

We sincerely thank all the users who contributed to the study. Also we would like to thank Marina Kovaleva for valuable discussions and Lana Kouchnir and Tatiana Logvinenko for help with manuscript preparation.

## References

An, S. M., Park, J. S., & Kim, S. H. (2014). Effect of energy drink dose on exercise capacity, heart rate recovery and heart rate variability after high-intensity exercise. Journal of Exercise Nutrition and Biochemistry, 18(1), 31–39. https://doi.org/10.5717/jenb.2014.18.1.31

Benichou, T., Pereira, B., Mermillod, M., Tauveron, I., Pfabigan, D., Maqdasy, S., & Dutheil, F. (2018). Heart rate variability in type 2 diabetes mellitus: A systematic review and meta–analysis. PLOS ONE, 13(4), e0195166. https://doi.org/10.1371/journal.pone.0195166

de Geus, E. J. C., Gianaros, P. J., Brindle, R. C., Jennings, J. R., & Berntson, G. G. (2019). Should heart rate variability be “corrected” for heart rate? Biological, quantitative, and interpretive considerations. Psychophysiology, 56(2), e13287. https://doi.org/10.1111/psyp.13287

Fu, Y., Huang, X., Piao, L., Lopatin, A. N., & Neubig, R. R. (2007). Endogenous RGS proteins modulate SA and AV nodal functions in isolated heart: Implications for sick sinus syndrome and AV block. American Journal of Physiology-Heart and Circulatory Physiology, 292(5), H2532–H2539. https://doi.org/10.1152/ajpheart.01391.2006

Garakani, A., Martinez, J. M., Aaronson, C. J., Voustianiouk, A., Kaufmann, H., & Gorman, J. M. (2009). Effect of medication and psychotherapy on heart rate variability in panic disorder. Depression and Anxiety, 26(3), 251–258. https://doi.org/10.1002/da.20533

Hasty, F., García, G., Dávila, H., Wittels, S. H., Hendricks, S., & Chong, S. (2021). Heart Rate Variability as a Possible Predictive Marker for Acute Inflammatory Response in COVID-19 Patients. Military Medicine, 186(1–2), e34–e38. https://doi.org/10.1093/milmed/usaa405

Hillebrand, S., Gast, K. B., de Mutsert, R., Swenne, C. A., Jukema, J. W., Middeldorp, S., Rosendaal, F. R., & Dekkers, O. M. (2013). Heart rate variability and first cardiovascular event in populations without known cardiovascular disease: Meta-analysis and dose–response meta-regression. EP Europace, 15(5), 742–749. https://doi.org/10.1093/europace/eus341

Hu, B., Guo, H., Zhou, P., & Shi, Z.-L. (2021). Characteristics of SARS-CoV-2 and COVID-19. Nature Reviews Microbiology, 19(3), 141–154. https://doi.org/10.1038/s41579-020-00459-7

Hughes, J. W., & Stoney, C. M. (2000). Depressed Mood Is Related to High-Frequency Heart Rate Variability During Stressors: Psychosomatic Medicine, 62(6), 796–803. https://doi.org/10.1097/00006842-200011000-00009

Huikuri, H. V., & Stein, P. K. (2013). Heart Rate Variability in Risk Stratification of Cardiac Patients. Progress in Cardiovascular Diseases, 56(2), 153–159. https://doi.org/10.1016/j.pcad.2013.07.003

Kim, H.-G., Cheon, E.-J., Bai, D.-S., Lee, Y. H., & Koo, B.-H. (2018). Stress and Heart Rate Variability: A Meta-Analysis and Review of the Literature. Psychiatry Investigation, 15(3), 235–245. https://doi.org/10.30773/pi.2017.08.17

Montano, N., Ruscone, T. G., Porta, A., Lombardi, F., Pagani, M., & Malliani, A. (1994). Power spectrum analysis of heart rate variability to assess the changes in sympathovagal balance during graded orthostatic tilt. Circulation, 90(4), 1826–1831. https://doi.org/10.1161/01.CIR.90.4.1826

Natarajan, A., Su, H.-W., & Heneghan, C. (2020). Assessment of physiological signs associated with COVID-19 measured using wearable devices. Npj Digital Medicine, 3(1), 156. https://doi.org/10.1038/s41746-020-00363-7

Quer, G., Radin, J. M., Gadaleta, M., Baca-Motes, K., Ariniello, L., Ramos, E., Kheterpal, V., Topol, E. J., & Steinhubl, S. R. (2021). Wearable sensor data and self-reported symptoms for COVID-19 detection. Nature Medicine, 27(1), 73–77. https://doi.org/10.1038/s41591-020-1123-x

Shaffer, F., & Ginsberg, J. P. (2017). An Overview of Heart Rate Variability Metrics and Norms. Frontiers in Public Health, 5, 258. https://doi.org/10.3389/fpubh.2017.00258

Taleghani, N., & Taghipour, F. (2021). Diagnosis of COVID-19 for controlling the pandemic: A review of the state-of-the-art. Biosensors and Bioelectronics, 174, 112830. https://doi.org/10.1016/j.bios.2020.112830

Tanwar, G., Chauhan, R., Singh, M., & Singh, D. (2020). Pre-Emption of Affliction Severity Using HRV Measurements from a Smart Wearable; Case-Study on SARS-Cov-2 Symptoms. Sensors, 20(24), 7068. https://doi.org/10.3390/s20247068

The, A.-F., Reijmerink, I., van der Laan, M., & Cnossen, F. (2020). Heart rate variability as a measure of mental stress in surgery: A systematic review. International Archives of Occupational and Environmental Health, 93(7), 805–821. https://doi.org/10.1007/s00420-020-01525-6

Tyapochkin, K., Smorodnikova, E., & Pravdin, P. (2019). Smartphone PPG: Signal processing, quality assessment, and impact on HRV parameters. 2019 41st Annual International Conference of the IEEE Engineering in Medicine and Biology Society (EMBC), 4237–4240. https://doi.org/10.1109/EMBC.2019.8856540

Zhou, F., Yu, T., Du, R., Fan, G., Liu, Y., Liu, Z., Xiang, J., Wang, Y., Song, B., Gu, X., Guan, L., Wei, Y., Li, H., Wu, X., Xu, J., Tu, S., Zhang, Y., Chen, H., & Cao, B. (2020). Clinical course and risk factors for mortality of adult inpatients with COVID-19 in Wuhan, China: A retrospective cohort study. The Lancet, 395(10229), 1054–1062. https://doi.org/10.1016/S0140-6736(20)30566-3

